# Correlations between *per capita* green tea consumption and COVID-19 morbidity/mortality: comparing the strength before and after Omicron emergence

**DOI:** 10.1101/2023.12.18.23300136

**Authors:** Maksim Storozhuk

## Abstract

**Purpose:** There is rapidly growing *in vitro* pharmacological evidence suggesting preventive/therapeutic potential of green tea and its constituent EGCG in battle with COVID-19. The aim of this study was to analyze time course of previously reported correlations between *per capita* green tea consumption and COVID-19 morbidity/mortality with a focus on comparing periods of time before (year 2021) and after (years 2022-2024) emergence of Omicron variant.

**Methods:** Correlations between *per capita* green tea consumption and COVID-19 morbidity/mortality were calculated using multiple regression models in a subset (n=84) of countries/territories worldwide with HDI above 0.55. Strength of the correlations for several dates in 2021 - 2024 was compared.

**Results:** Higher *per capita* green tea consumption was associated with lower COVID-19 morbidity and mortality. Statistically significant correlations were observed in 2021, 2022, 2023 and 2024. As compared to 2021, the strength of both correlations was decreased during 2022-2024 period. On average normalized to 2021 strength of correlation between *per capita* green tea consumption and COVID-19 morbidity decreased to 0.60, while normalized strength of correlation between *per capita* green tea consumption and COVID-19 mortality decreased to 0.83.

**Conclusion:** Overall, the results of this study are in line with *in vitro* pharmacological evidence. Though indirectly, these results at epidemiological level support the idea that green tea consumption may have not only preventive, but also therapeutic value in relation to COVID-19. Nevertheless, because of limitations of this study, this idea still should be considered as a hypothesis requiring further assessment.

## Introduction

In spite of the development of numerous vaccines for the prevention of COVID-19 and approval of several drugs for its treatment, there is still a great need for effective and inexpensive therapy of this disease. There is rapidly growing *in vitro* pharmacological evidence suggesting preventive/therapeutic potential of green tea and its constituent EGCG in battle with COVID-19 (see [1-3] for review). There is also evidence supporting this idea in an animal model [4]. At least three molecular targets of EGCG in relation to SARS-CoV-2 have been identified (e.g. see [3] for a review). At the same, observational studies accessing this issue are scarce (but see [5], results of relevant controlled clinical studies have not yet been reported, though results of proof-of-principle study [6] are promising. Thus, addressing this issue using an ecological approach seems to be reasonable in spite of its limitations [7, 8]. Using this approach we have recently reported that correlations between *per capita* green tea consumption and COVID-19 morbidity/mortality are still observed after emergence of Omicron variant, and this is in line with our pharmacological evidence showing that green tea and EGCG inhibit the activity of the omicron variant 3CL protease (enzyme essential for virus replication) efficiently [9]. At the same time we noted possible decrease of strength of the abovementioned correlations, as compared to period of time before Omicron emergence [9]. Here this possible decrease was examined in more detail. Apart from other reasons, this examination was encouraged by recent report of Shin-Ya and colleagues [10]. In particular, they showed that: (i) EGCG inhibits interaction between receptor binding domain (RBD) of S-protein of Omicron (BA.1) with ACE2, and this reduces the infectivity of the virus; (ii) EGCG is substantially less effective in reducing infectivity of *some* sub-variants of Omicron. Thus, it cannot be excluded that our observation regarding the decrease of strength of the correlations (given it is confirmed) reflects the decreased effect of EGCG on interaction between RBD and ACE2.

## Methods

### Ethics approval and consent to participate

Not applicable (ethical approval was not deemed necessary as this was an analysis of publicly available data).

All data were obtained from open sources. Information about COVID-19 morbidity (defined as total number of cases per million population) and mortality (defined as total number of deaths per million population) for a specific date was directly obtained from ‘Worldometers info. Coronavirus’ [11]. The information on ‘Worldometer’ is based on official reports and considered as a reliable (for instance, [12]. Analysis used in this report was similar to that described previously [7-9]. However, here the focus was on the strength of correlations between *per capita* green tea consumption and COVID-19 morbidity/mortality in a subset of countries/territories with HDI above 0.55 (n=84).

Correlations between *per capita* green tea consumption and COVID-19 morbidity (or mortality) were calculated using multiple regression models accounting for several confounding factors. Specifically, beside *per capita* green tea consumption the following factors were included: population density; percentage of population aged above 65; percentage of urban population; Human Developmental Index (HDI). In a complementary analysis an additional variable-vaccination rates was added.

It should be noted that while strength of correlations between *per capita* green tea consumption and COVID-19 morbidity/mortality have been already reported for several dates before Omicron emergence [7, 8], these values cannot be *directly* compared to values reported more recently [9]. Indeed, subsets of countries used for multiple linear regression analysis were different: countries with HDI above 0.55 only [9], versus countries with HDI above and below 0.55 [7, 8]. Additionally, logarithm of per capita green tea consumption was used in multiple linear regression model in more recent work [9] while per capita green tea consumption itself was used in earlier works [7, 8]. (Variables COVID-19 morbidity and COVID-19 mortality) were transformed to Log in all abovementioned ecological studies).

Unless otherwise noted COVID-19 morbidity and COVID-19 mortality referred in this report reflect *cumulative* data since the beginning of the epidemic to specified date. Though *increases* in COVID-19 morbidity and mortality for specified periods of time were also calculated (as described earlier [9] see also Supplementary Materials), this was done in complementary analysis to access possible bias of results due to vaccination.

KyPlot’ software was used for statistical analysis.

## Results

As already mentioned, here the time course of correlations between *per capita* green tea consumption and COVID-19 morbidity/mortality was analyzed with a focus on comparison periods of time before and after emergence of Omicron variant. In particular, results of the comparisons are shown in **Fig. 1**. First of all it is interesting to mention that, correlation between *per capita* green tea consumption and COVID-19 mortality is slightly but systematically stronger than that between *per capita* green tea consumption and COVID-19 morbidity. Additionally, levels of statistical significance of correlations between *per capita* green tea consumption and COVID-19 mortality are higher for most of the accessed dates (**Fig 1**). While both correlations are still statistically significant during 2022-2024 period, the strength of both correlations appears to be weaker as compared to year 2021.

**Figure 1.**
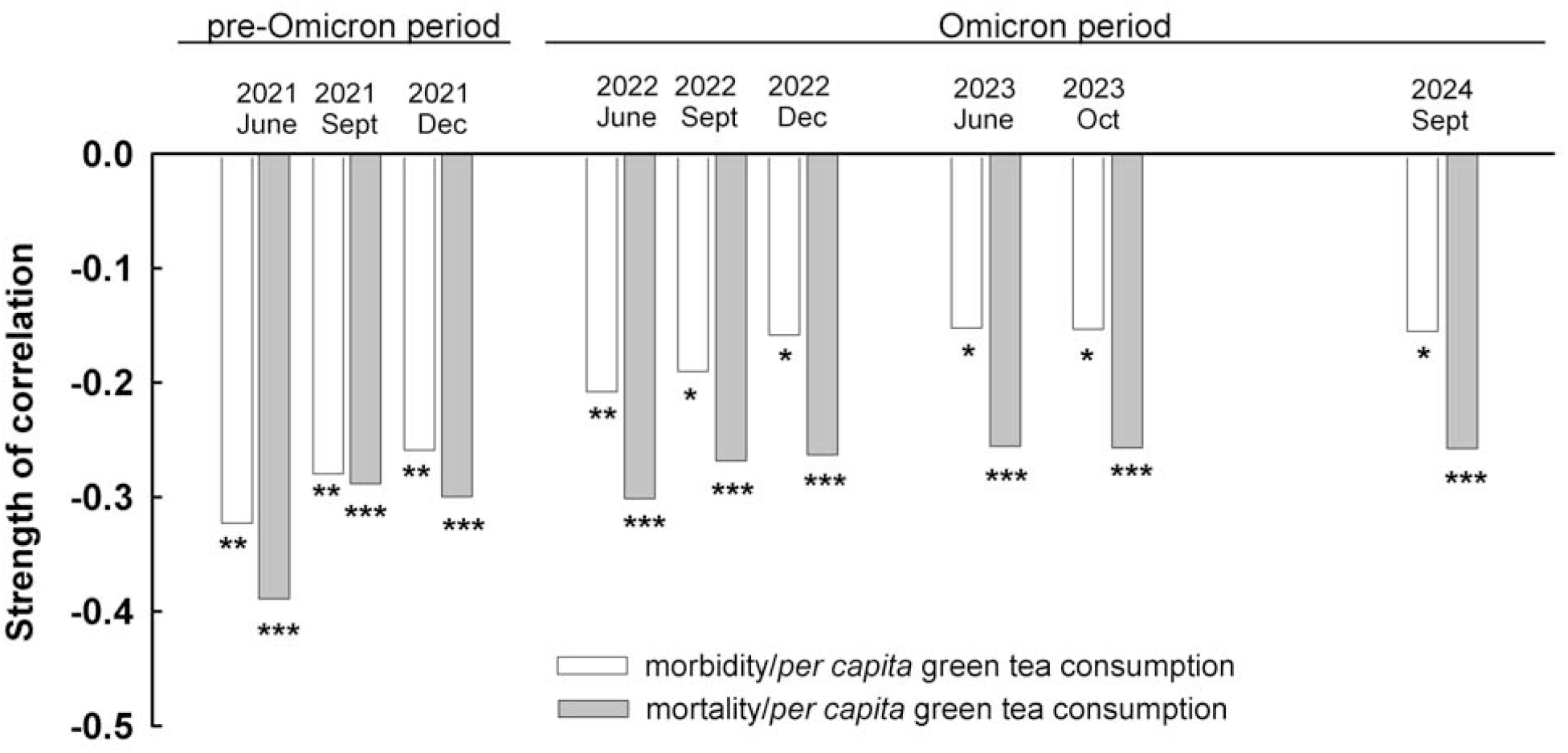
Strength of correlation between per capita green tea consumption and COVID-19 morbidity/mortality: changes over time. The values are coefficients of regression obtained in the multiple regression models accounting for the following confounding factors: population density, percentage of population aged above 65, percentage of urban population, and Human Developmental Index (HDI). See methods and [9] for more details. *, **, and *** indicate levels of statistical significance of these correlations: P<0.05, P<0.01 and P<0.001 respectively.

At the same time the *relative* decrease of the strength of correlation between *per capita* green tea consumption and COVID-19 mortality seem to be notably and systematically lower for all accessed dates during years 2022 and 2023 (**Fig 2 A**). Indeed, as compared to year 2021, on average normalized strength of correlation between *per capita* green tea consumption and COVID-19 morbidity decreased to 0.60± 0.032, while normalized strength of correlation between *per capita* green tea consumption and COVID-19 morbidity decreased to 0.83±0.022 (**Fig 2 B**). The decrease of the strength of correlation between *per capita* green tea consumption and COVID-19 morbidity was statistically significant (P= 0.02, Mann-Whitney U Test for Unpaired Data), while the decrease in strength of correlation between *per capita* green tea consumption and COVID-19 morbidity was not (P= 0.07 Mann-Whitney U Test for Unpaired Data).

**Figure 2.**
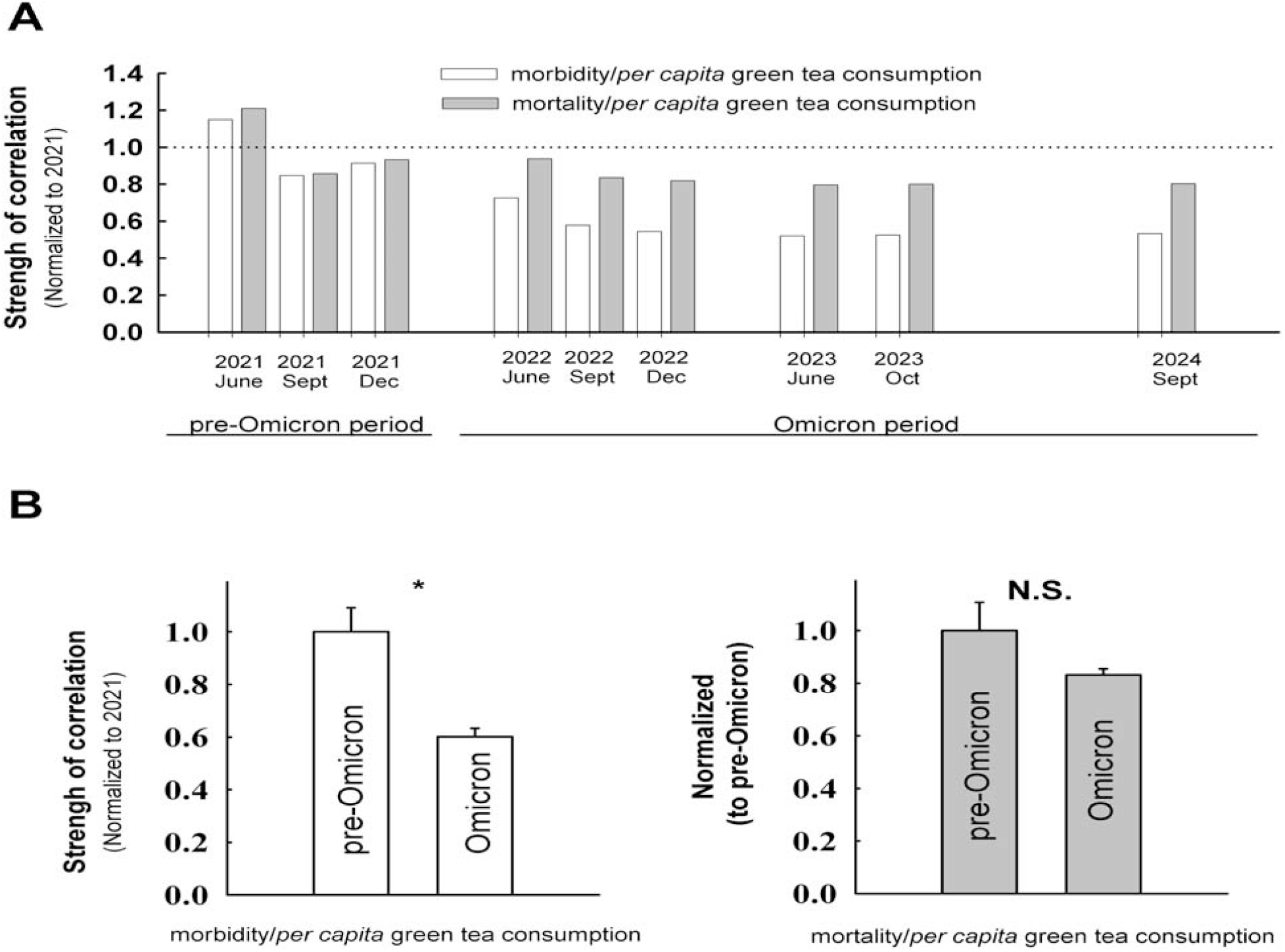
Differential decrease of strength of correlations between per capita green tea consumption and COVID-19 morbidity/mortality during Omicron period. A. Same data as in **Fig.1** normalized to average of the values in the year 2021. B. Summary for normalized data. Notice more pronounced decrease of strength of correlation between per capita green tea consumption and COVID-19 morbidity. Data presented as mean ±S.E.M. * - indicates statistically significant difference between columns (P<0.05).

Altogether the results above hint that the correlation between *per capita* green tea consumption and COVID-19 mortality reflects *not only* corresponding correlation with the morbidity, but also some other factors/mechanisms. In particular, based on these epidemiological results, it may be expected that green tea consumption may have not only preventive, but also therapeutic effects *in vivo*. Overall this expectation is in line with *in vitro* pharmacological evidence indicating that green tea constituents affect separate molecular targets responsible for: (i) entry of the virus; (ii) virus replication.

It should be noted that rates of vaccination against COVID-19 were not included in linear models described above. Potentially, this may bias the results. To address this concern additional analysis was performed. In this analysis *increases* in COVID-19 morbidity and mortality for specified periods of time were calculated. Strength of correlations between *per capita* green tea consumption and COVID-19 morbidity /mortality was compared in models: (i) not including vaccination rates as *additional* confounding factor; and (ii) including this factor (vaccination rates in the middle of corresponding period). See Methods and Supplementary materials for more details. This approach was applied to periods December 14, 2020 - December 6, 2021 and December 6, 2021 - December 6, 2022 (considered as mostly pre-Omicron and Omicron, respectively). Inclusion of vaccination rates as additional confounding factor in models did not substantially affected strength of the correlations considered in this report. Indeed, strength of the correlations was nearly the same with and without adjustment for vaccination rates **Fig. 3 (A, B)**.

**Figure 3.**
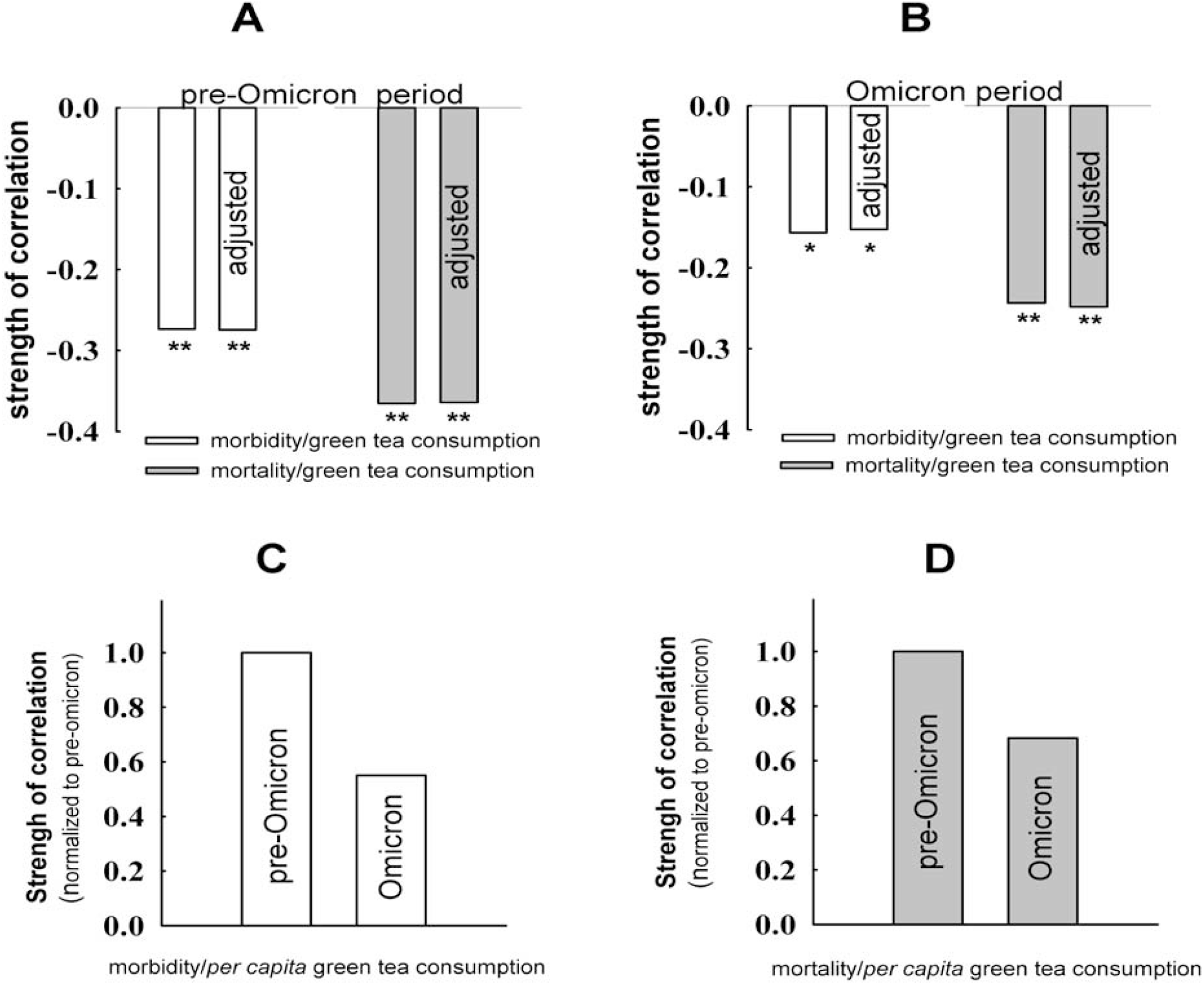
Analysis of increases in COVID-19 morbidity/mortality during specified periods. **(A, B)** adjustment for vaccination rates does not affect strength of correlations between per capita green tea consumption and COVID-19 morbidity/mortality. **(C, D)** Comparing relative decreases of strength of correlations between per capita green tea consumption.

**Fig. 3** also illustrates that during Omicron period strength of correlations between *per capita* green tea consumption and COVID-19 morbidity/mortality is decreased as compared to pre-Omicron period. This is in line with results shown in **Fig. 1**. Quantitative differences are not surprising, considering that **Fig. 1** considers cumulative morbidity/mortality since beginning of epidemic, while **Fig. 2 -** considers increases during specified periods.

## Discussion

In spite of the development of numerous vaccines for the prevention of COVID-19 and approval of several drugs for its treatment, there is still a great need for effective and inexpensive therapy of this disease. Additionally, it should be mentioned that initial expectations regarding efficacy and safety of SARS-CoV-2 vaccines were, probably, exaggerated (see [13] for review). In this overall context, green tea catechins, are considered as promising because of their antiviral effects against SARS-CoV-2, while EGCG was reported as most effective (e.g. see [1-3] for review). There is ongoing discussion regarding relative contribution of distinct molecular targets of EGCG in its overall effects in relation to COVID-19. Some groups propose that effects of EGCG in relation to COVID-19 are mostly if not exclusively due to inhibition of interaction between RBD of S-protein of SARS-CoV-2 with ACE2 in host cells, and the effects are predominantly/exclusively preventive. Other groups propose that effects of EGCG on 3CL protease are of primary importance, considering that inhibition of 3CL protease prevents replication of the virus, therapeutic effects of EGCG should be expected. Both viewpoints are supported by solid *in vitro* pharmacological evidence and, in some cases, evidence in animal models (see [2, 3] for review). However, there is much less clarity regarding applicability of these important finding in actual fight with COVID-19 as outlined in questions below. Can at least one of molecular targets of green tea catechins be affected by green tea consumption *per ce*? Or, only a special formulation, with increased concentrations of green tea catechins/(additives enhancing bioavailability of green tea catechins), must be used for this purpose? Would such a formulation be efficient and safe at the same time? Direct answers to these questions can be obtained only in clinical trials, preferably, large-scale, randomized and placebo controlled. Considering numerous reasons (e.g., ethical, complexity of design and high costs of relevant clinical trials), it is not surprising that results of such trials have not been reported, even in relation to pre-Omicron variants of SARS-CoV-2. Therefore, indirect evidence addressing these issues should be considered as potentially important. Below results obtained in the current report will be discussed in relation to the first of above-mentioned questions. The focus will be on possible similarities or differences as compared to results obtained using pharmacological approach.

Basically, results obtained in this report can be summarized as follows.

1. Higher *per capita* green tea consumption is associated with lower COVID-19 morbidity and mortality.
2. Correlations between *per capita* green tea consumption and COVID-19 morbidity/mortality are still statistically significant during Omicron period.
3. Strength of both correlations is decreased during Omicron period.
4. At the same time relative decrease of strength of correlation between *per capita* green tea consumption and COVID-19 morbidity (∼40%) is more pronounced than decrease of strength of correlation between *per capita* green tea consumption and COVID-19 mortality (∼20%).

1. Results obtained in this report confirm and extend previous [7]-9] observations. Higher *per capita* green tea consumption is associated with lower COVID-19 morbidity and mortality. Statistically significant correlations were observed during 2021-2024 period. Due to limitations of ecological approach these results, taken alone, could not prove a causal relation. On the other hand, possibility that this relation is causal is supported by results obtained using other approaches. Indeed, there is mounting evidence indicating that apart of beneficial effects of green tea constituents in relation to comorbidities associated with COVID-19 (e.g., [14-17]), some of these constituents (EGCG is considered as most effective) *directly* interact with several proteins essential for functioning of SARS-CoV-2 (see [3] for review). Indeed, apart from numerous *in silico* studies (e.g. [18, 19]) this is supported by rapidly growing *in vitro* pharmacological evidence ((see [1-3] for review). There is also important relevant evidence obtained in an animal model [4], and in studies showing that effects of green tea constituents on infectivity of SARS-CoV-2 are still observed in *saliva* of healthy volunteers [20, 10]. The latter is rather supported by results of an observational study reporting that people who consumed ≥4 cups/day of green tea had a lower, albeit statistically not significant, odds of SARS-CoV-2 infection [5]. In the discussion authors suggest that “present study was not large enough to detect the observed association with statistical significance” [5]. Unfortunately, this seems to be the only observational study addressing this issue. Regarding future observational studies and clinical trials addressing this issue, it should be pointed out that consumption of green tea beverages by participants should be accounted for. Indeed, green tea beverages contain substantial amounts of green tea catechins and have pronounced effects on SARS-CoV-2 [21]. Otherwise, results can be biased towards *underestimation* of the effects, especially in countries where green tea beverages are popular. It should be mentioned that even after regular tea drinking, concentrations of EGCG, at least in some parts of the upper respiratory tract, are high enough to substantially inactivate known molecular targets of SARS-CoV-2 (see [3] for detailed review). Moreover, these high concentrations persist at least minutes *after* green tea consumption [3] (or a beverage containing green tea catechins [22]). Thus, at least preventive effects of green tea in relation to COVID-19 should not be considered as surprising.
2. Correlations between *per capita* green tea consumption and COVID-19 mortality/morbidity were still statistically significant after Omicron emergence (2022-2023). This is well in line with pharmacological evidence obtained *in vitro*. Indeed, EGCG as well as green tea inhibit the activity of the *Omicron* variant 3CL protease efficiently [9]. It was also shown that EGCG and green tea affect RBD of S-protein of Omicron variant and decrease infectivity of the virus [10]. Thus, at least two different targets of EGCG in relation to COVID-19 are strongly affected in both, pre-Omicron and Omicron variants.
3. Strength of both correlations is decreased during omicron period. Is there pharmacological evidence indicating decreased effects of green tea constituents on respective molecular targets of Omicron as compared to previous variants of SARS-CoV-2? In respect of 3CL protease it was reported that effects of EGCG on WT- and *Omicron-* 3CL proteases are comparable [9]. Results of similar comparisons regarding effects of EGCG on RBD of S-protein of Omicron versus that of WT (or some pre-Omicron variants) were not. Nevertheless, as outlined below, a tentative answer is that effects of EGCG on RBD of S-protein of Omicron are weaker as compared to WT/pre-Omicron variants. Infectivity of WT/pre-Omicron variants ((UK-B.1.1.7, SA-B.1.351, and CA-B.1.429), was reduced *at least* 10 times by 100 μM EGCG ((see Fig. 3 (C,D) in [23]). Indeed, EGCG (100 μM) reduced the infectivity to less than 5% (WT, SA-B.1.351, and CA-B.1.429) or 10 % (B.1.1.7). On the other hand, at concentration 125 μM, EGCG reduced the infectivity of Omicron variant (BA.1, BA.2, BA.5, BQ.1.1, XBB.1, BA.2.75 and XE) less than 10 times (see Figure 2. in [10]. Another figure there (Figure 4. in [10]) gives essentially the same hint: EGCG (125) μM reduces infectivity of BA.1 sub-variant of Omicron by ∼70 %. Moreover, as compared to BA.1, effects of EGCG on infectivity of several other Omicron sub-variants (BA.5, BA.2.75 and BQ.1.1) were strongly reduced and this is emphasized by authors [10]. Thus, in respect of effects of EGCG on RBD of S-protein, it is likely that “average” effects of EGCG on Omicron variant are reduced, as compared to “average” effects on pre-Omicron variants. Nevertheless, a direct answer to this question ((i.e. comparison in study with exactly the same experimental protocol) seems to be interesting and potentially important. For instance, if the decreased effects of EGCG on Omicron variant will be confirmed, it will be additional argument supporting: (i) importance of EGCG influence on RBD of S-protein *in vivo*; (ii) causal link between *per capita* green tea consumption and COVID-19 mortality/morbidity
4. As already mentioned, the strength of both correlations considered in this report was decreased during Omicron period. At the same time the decrease of strength of correlation between *per capita* green tea consumption and COVID-19 morbidity (∼40%) appear to be more pronounced than the decrease of strength of correlation between *per capita* green tea consumption and COVID-19 mortality (∼20%) (**Figure 2)**. Additionally, levels of statistical significance of correlations between *per capita* green tea consumption and COVID-19 mortality are higher for most of the accessed dates (**Fig 1**). Together these results suggest that the correlation between *per capita* green tea consumption and COVID-19 mortality reflects *not only* corresponding correlation with the morbidity, but also some other factors/mechanisms. These results may be of relevance to ongoing discussion regarding relative contribution of distinct molecular targets to overall effects of green tea catechins on SARS-CoV-2 (preventive via RBD of S-protein versus therapeutic via 3CL protease). Based on these epidemiological results, it may be expected that green tea consumption may have not only preventive, but also therapeutic effects *in vivo*. Overall this expectation is in line with *in vitro* pharmacological evidence indicating that green tea constituents affect separate molecular targets responsible for: (i) entry of the virus; (ii) virus replication.

### Conclusion

Overall, the results of this study are in line with *in vitro* pharmacological evidence. Though indirectly, these results at epidemiological level support the idea that green tea consumption may have not only preventive, but also therapeutic value in relation to COVID-19. Nevertheless, because of limitations of ecological approach, this idea still should be considered as a hypothesis requiring further assessment.

## Declarations section

### Ethical Approval and Consent to participate

Not applicable

## Consent for publication

Not applicable

## Availability of supporting data

Data (mostly) available within the article or its supplementary materials. Additional data available on request from the author.

## Acknowledgements

I would like express my gratitude to personnel of the open resources for creating excellent opportunities for research.

## Supplementary materials

### Methods in more details

#### Data regarding COVID-19 morbidity and mortality

As already mentioned, the methodological approach used in this report is similar to that described previously [7-9]. Information about COVID-19 morbidity (defined as total number of cases per million population) and mortality (defined as total number of deaths per million population) for a specific date was directly obtained from ‘Worldometers info. Coronavirus.’ The information on ‘Worldometer’ is based on official daily reports and considered reliable [12;24]. Analysis was restricted to countries or territories (according UN classification): with a population of at leas 3 million; listed on ‘Worldometers info’; known estimate of *per capita* green tea consumption; HDI above 0.55. Thus, analysis was restricted to 84 countries/ territories.

Unless otherwise noted COVID-19 morbidity and COVID-19 mortality referred in this report reflect *cumulative* data since the beginning of the epidemic to specified date (e.g., data shown in Figures 1 and 2 reflect epidemiological situation as of: June 3, 2021; September 15, 2021, December 6, 2021; June 2, 2022; September 15, 2022; December 6, 2022; June 25, 2023, October 22, 2023, September 15, 2024). However, *increases* in COVID-19 morbidity and mortality for specified period of time were also calculated. Specifically, increases for two periods of time were calculated: (i) December 14 - 2020 December 6, 2021; (ii) December 6, 2021 - December 6, 2022. In this analysis differences in total number of cases/deaths were calculated first, and then these values were divided by country population (in millions). These specific periods for analysis were chosen based on following: 1) the periods should reflect epidemiological situation (mostly) before and after Omicron emergence, respectively; 2) availability of archived data tables regarding the cases/deaths; 3) periods should be close to a calendar year to minimize seasonal variations in morbidity and mortality (to reduce potential bias to this factor).

#### Statistical analysis

Since the variables of COVID-19 morbidity and COVID-19 mortality do not have a normal distribution [12], a non–parametric statistic was primarily employed for the analysis as suggested earlier [12]. For multiple linear regression analysis, an approach similar to that previously reported [12] was used. In this analysis, morbidity and mortality per million of the population were transformed into common logarithm (log10) to adjust for normality of the distribution, as suggested previously [12]. For the same reason *per capita* green tea consumption was also transformed into common logarithm (log10), which notably decreased skewness of the distribution of this variable. Logarithmic dependence between dose and effect, usually observed in pharmacological studies, was an additional argument to use this transformed variable in multiple linear regression models. Other factors included in multiple linear regression analysis, were population density, percentage of population aged above 65, percentage of urban population, and Human Developmental Index (HDI), which is based on access to health and education services and income [25]. In the analysis of specified periods (December 14, 2020 - December 6, 2021 and December 6, 2021 - December 6, 2022), vaccination rates were *also* included in the linear regression model. The values were retrieved from Online Resource OurWorldInData.org (https://ourworldindata.org/coronavirus). Specifically, values corresponding to the middle of the periods analyzed in this study were used for analysis.

‘KyPlot’ software was employed for statistical assessments.

